# Molecular & Translational Biology of the Blood-Based VeriStrat® Proteomic Test Used in Cancer Immunotherapy Treatment Guidance

**DOI:** 10.1101/2022.12.28.22283689

**Authors:** Matthew A Koc, Timothy Aaron Wiles, Daniel C Weinhold, Steven Rightmyer, Joanna Roder, Senait Asmellash, Heinrich Roder, Robert W Georgantas

## Abstract

**INTRODUCTION:** The blood-based VeriStrat^®^ proteomic test (VS) predicts patient response to therapy based on the intensities of eight different features in a mass spectrum obtained from MALDI-TOF analysis of human serum/plasma specimens. An interim analysis of the INSIGHT clinical trial (NCT03289780) demonstrated that VS labels, VS Good and VS Poor, predict patients with non-small cell lung cancer (NSCLC) who are likely sensitive or resistant to immune checkpoint inhibitor (ICI) therapy [1]. While VS measures intensities of eight spectral features by matrix-assisted laser desorption/ionization (MALDI) time-of-flight (TOF) mass spectrometry from patient serum/plasma samples, the individual proteoforms underlying these features have not been rigorously and comprehensively identified.

**OBJECTIVES:** The objective of this study was to identify the proteoforms measured by VS.

**METHODS:** Mass spectra for VS are acquired using a standard low-resolution MALDI-TOF procedure that generates broad, composite features. DeepMALDI [2] analysis of serum samples was used to resolve these features into finer peaks. Top-down proteomics analysis of human serum, combining reversed-phase fractionation and liquid chromatography – tandem mass spectrometry (LC-MS/MS), was then used to identify the key proteoform constituents of these peaks.

**RESULTS:** It was determined that proteoforms of serum amyloid A1, serum amyloid A2, serum amyloid A4, C-reactive protein, and beta-2 microglobulin are primary constituents of the VS spectral features.

**CONCLUSION:** Proteoforms of several proteins related to host immunity were identified as major constituents of these features. This information advances our understanding of how VS can predict patient response to therapy and opens the way for further translational studies.

**Highlights:**

- The combination of top-down proteomics and DeepMALDI^®^ spectrometry enables the identification of proteoforms measured by the VeriStrat Proteomic test.
- Proteoforms of serum amyloid A1 (SAA1), SAA2, SAA4, beta-2 microglobulin, and C-reactive protein are the primary constituents of the spectral features measured in the VeriStrat proteomic test.
- The proteins assayed by the VeriStrat proteomic test have individual prognostic value for oncology and immuno-oncology outcomes.
- The proteins assessed by the VeriStrat proteomic test have been shown to have direct effects on patient immune activity.

## Introduction

The VeriStrat (VS) test is a mass spectrometry-based, proteomic assay that is performed on human serum and plasma. Details of the development and initial validation of the test have been published elsewhere [3]. Briefly, mass spectra are acquired in triplicate from the patient sample using a matrix-assisted laser desorption/ionization (MALDI) time-of-flight (TOF) mass spectrometer. The spectra are processed to render them comparable between samples by background subtraction, alignment and normalization, and the area under the processed spectra in eight mass spectral regions is evaluated. The definitions of these eight spectral features, VS1-VS8, are listed in Table 1.

**Table 1.** Definitions of the mass spectral features used in the VS classification algorithm.

| Feature Name | Left boundary m/z | Right boundary m/z |
| --- | --- | --- |
| VS1 | 5,811.09 | 5,875.39 |
| VS2 | 11,376.15 | 11,515.35 |
| VS3 | 11,459.32 | 11,599.72 |
| VS4 | 11,614.02 | 11,756.72 |
| VS5 | 11,687.26 | 11,831.06 |
| VS6 | 11,830.29 | 11,976.19 |
| VS7 | 12,375.38 | 12,529.38 |
| VS8 | 12,501.85 | 12,657.85 |

The values of these eight mass spectral features are compared with those of the samples in the classification-labelled reference set using a 7-nearest neighbor algorithm to yield a “VS Good” or “VS Poor” determination for each spectrum. If the three replicate spectra generated from a sample generate concordant results, i.e., all “VS Good” or all “VS Poor”, that result is returned as the VS classification. If the replicate results are discordant, an “Indeterminate” classification is reported. Spectral quality is ensured via multiple quality control checks. For example, the spectra must contain a minimum number of detectable peaks, the noise level must not exceed a maximum limit, and a minimum number of peaks must be used during spectral alignment. The spectra are also monitored to ensure that a VS Poor result is not generated via existence of a large interfering peak at m/z = 11.7 kDa, which is rarely observed, due to extremely high levels of beta-2 microglobulin (B2M), such as in patients with severely compromised liver function. [4, 5]

The abundances of multiple serum proteins are correlated with VS classification, including C-reactive protein (CRP), interleukin-6 (IL-6), serum amyloid A (SAA), the cytokeratin 19 fragment CYFRA 21-1, insulin-like growth factor 2 (IGF-2), and osteopontin [6]. Set enrichment analysis techniques have shown that VS classification is associated with acute phase response [6-8], acute inflammatory response, and complement activation [7, 8]. Indeed, it is well known that truncated proteoforms of serum amyloid A are present in the mass/charge range of some of the VS features in Table 1 [9, 10].

The VS test has demonstrated both prognostic and predictive power in the advanced non-small cell lung cancer (NSCLC) setting and in other tumor types. The ability of the test to predict differential benefit from erlotinib vs single-agent chemotherapy between patients with advanced NSCLC assigned a VS Poor or VS Good classification of pretreatment serum was demonstrated in a prospective, randomized clinical trial [11]. Individual studies and meta-analysis have shown that patients assigned a pretreatment VS Good classification have significantly better outcomes than patients assigned a VS Poor classification across multiple treatment regimens and in the absence of active therapy in the advanced NSCLC setting [12-15]. Most recently, the VS test has shown potential as a prognostic marker for immunotherapy treatment of advanced NSCLC [1, 16, 17]. The test may also have utility in guiding treatment by predicting those patients who obtain most benefit from the addition of chemotherapy to ICI [1]. The prognostic power of the test has been identified in other advanced solid tumors, including colorectal cancer, squamous cell cancer of the head and neck [18], pancreatic cancer [19], and breast cancer [20].

The current study was undertaken to definitively determine the protein constituents of the VS features, with a translational biology goal of understanding how those proteins may play a role in immuno-oncology. The study also investigated why the presence of these proteins detected in the VeriStrat proteomic test may impart its ability to predict NSCLC immunotherapy outcomes.

## Material and Methods

### Samples

Serum samples from human subjects diagnosed with lung cancer were acquired from commercial biobanks (Discovery Life Sciences (Huntsville, AL), ProMedDx (Norton, MA), ProteoGenex (Inglewood, CA) and BioIVT (Westbury, NY)) or from patients enrolled in the PROSE study (Randomized Proteomic Stratified Phase III Study of Second-Line Erlotinib Versus Chemotherapy in Patients With Inoperable Non-Small Cell Lung Cancer, NCT00989690). All samples were collected under ethics-approved protocols according to Institutional Review Board/Independent Ethics Committee requirements.

Of 53 serum samples purchased from the biobanks, 26 serum samples with VS good status and 16 serum samples with VS poor status were combined to create large pools of VS Good and VS Poor serum, respectively. The pooled samples are referred to as VSP and VSG hereafter. The pooled samples were used for top-down mass spectrometry analysis and immunodepletion of serum amyloid A and C-reactive protein. Two additional samples with VS Good and VS Poor status, called VS Good and VS Poor, were not included in the VSG and VSP pools.

We also used 199 remnant samples from the PROSE study [11] for DeepMALDI [2] analysis.

### Reagents

Antibodies: Biotin-conjugated mouse monoclonal anti-human antibody to serum amyloid A (SAA; Cat. No. MO-C40028TB) and C-reactive protein (CRP; Cat. No. MAB5101) were purchased from Anogen (Mississauga, ON, Canada,) and VWR International (Radnor, PA), respectively.

Solvents and other materials: Acetonitrile, HPLC grade water, and trifluoroacetic acid were purchased from VWR International (Radnor, PA). Phosphate-buffered saline (PBS) and Dynabeads™ M-280 streptavidin were purchased from ThermoFisher Scientific (Carlsbad, CA). Bondapak^®^ C18 bulk packing material (125 Å pore size, 37-55 μm particle diameter) was purchased from Waters (Milford, MA). Sinapinic acid was purchased from Proteochem (Loves Park, IL). Serum cards were purchased from GE HealthCare (Buckinghamshire, UK). PBS with 0.05% Tween^®^ 20 (PBST) was purchased from Millipore Sigma (Burlington, MA). Protein Calibration Standard I was purchased from Bruker (Billerica, MA).

### VS spectral acquisition

VS spectra were acquired on a microflex^®^ MALDI-TOF mass spectrometer (Bruker, Billerica, MA) equipped with a 337 nm nitrogen laser with a maximum repetition rate of 60 Hz. The instrument was operated in positive-ion, linear mode, scanning a mass range of m/z 3000 to 30,000 Da. The 500 shot spectra (in 25 shot steps) were acquired from 3 replicate spots per sample after external calibration of the instrument with a mixture of calibrants in Protein Calibration Standard I. An average spectrum of 1500 shots was generated for each serum sample analyzed.

Serum samples for VS spectra acquisition were prepared as follows. Serum samples were thawed and 10 μl aliquots of each sample were spotted onto serum cards. The cards were allowed to dry for 1 hour at ambient temperature after which three 3 mm punches were excised from the serum spot. All three punches were placed in a 1.5 mL tube to which 50 μL of water was added. The punches were vortexed gently for 3 minutes. Twenty microliters of the serum extract were transferred to a 0.5 mL tube. Twenty microliters of matrix (25 mg of sinapinic acid per 1 mL of 50% acetonitrile:50% water plus 0.1% trifluoroacetic acid; percentages are volume/volume) were added to 20 μL of serum extract and mixed by vortexing. Three aliquots of 2 μL sample:matrix mix were then spotted onto an MALDI target (Hudson Surface Technology, Closter, NJ). The MALDI target was allowed to dry on the bench before placement in the MALDI mass spectrometer.

VS spectra were analyzed according to the standard protocols used in clinical specimen testing for VeriStrat.

### DeepMALDI spectral acquisition

DeepMALDI [2] spectra were acquired on a rapifleX^®^ MALDI-TOF mass spectrometer (Bruker, Billerica, MA) equipped with a Bruker smartbeam™ laser. The instrument was operated in positive-ion, linear mode, scanning a mass range of m/z 3000 to 30,000 Da. External calibration was performed using peaks m/z = 3320.0000, 4158.7338, 6636.7971, 9429.3020, 13,890.4398, 15,877.5801, and 28,093.9510 or Bruker’s Protein Calibration Standard I, consisting of a mixture of insulin, ubiquitin I, cytochrome C, and myoglobin.

Serum samples were prepared for DeepMALDI analysis as follows. Serum samples were thawed and 3 μL aliquots of each sample were spotted onto serum cards. The cards were allowed to dry for 1 hour at ambient temperature after which the whole serum spot was punched out with a 6 mm skin biopsy punch. Each punch was placed in a centrifugal filter with 0.45 μm nylon membrane. One hundred microliters of HPLC grade water were added to the centrifugal filter containing the punch. The punches were vortexed gently for 10 minutes then spun down at 14,000 rcf for 2 minutes. The flow-through was removed and transferred back on to the punch for a second round of extraction. Twenty microliters of the filtrate from each sample were then transferred to a 0.5 mL microcentrifuge tube for MALDI analysis. All subsequent sample preparation steps were carried out in a custom-designed humidity and temperature control chamber (Coy Lab Products, Grass Lake, MI). The temperature was set to 30 °C and the relative humidity to 10%.

An equal volume of freshly prepared matrix (25 mg of sinapinic acid per 1 mL of 50% acetonitrile:50% water plus 0.1% trifluoroacetic acid) was added to each 20 μL serum extract and the mix vortexed for 30 seconds. The first five aliquots (5 × 2 μL) of sample:matrix mix were discarded. Eight aliquots of 2 μL sample:matrix mix were then spotted onto a stainless steel MALDI target plate (Bruker, Billerica, MA).

For immunodepletion experiments, the reconstituted eluents from the immunodepleted serum samples were mixed with an equal volume of matrix solution (25 mg of sinapinic acid per 1 mL of 50% acetonitrile:50% water plus 0.1% trifluoroacetic acid) and spotted onto stainless steel MALDI plates for spectra acquisition on the Bruker rapifleX.

The MALDI target was allowed to dry in the environmental control chamber before placement in the MALDI mass spectrometer.

DeepMALDI spectra were analyzed using in-house software developed to deal with the large dynamic range observed in serum [21]. In particular, for quantitative analysis of the spectra in the PROSE set, only the strongly background-subtracted peak structure denoted as ‘FINE’ in [21] were used. This mitigates tails of large peaks from contributing to the intensity of smaller neighboring peaks. After background subtraction, peak intensities were normalized across samples using the intensity of peaks with the smallest variation across samples (as determined by an iterative process). Peaks used for normalization were at m/z: 3433, 3556, 3564, 3684, 3711, 3782, 3831, 4017, 4578, 4604, 4693, 5116, 5415, 6206, 6541, 7054, 7064, 8255, 9724, 9737, 14039, 14054, 14070, 14101, 14118, 14141, 14159, 14178, 14261, 14273, 14395, and 28102.

### Calibration of MALDI-TOF m/z axis

To ensure mass accuracy of DeepMALDI spectra, the Bruker rapifleX was calibrated using Bruker’s Protein Calibration Standard I – a mixture of insulin, ubiquitin I, cytochrome C, and myoglobin – and reference spectra were acquired. The m/z axis of all DeepMALDI spectra were then aligned to these reference spectra. Accuracy of the aligned m/z axes was confirmed by evaluation of the peaks corresponding to known proteoforms within the m/z range of the calibration standard but that are located outside of VS features.

### Preparation of samples for top-down proteomics

To prepare serum samples for top-down proteomics analysis, 50 μL of serum were diluted with 50 μL of water and filtered through a 0.2 μm modified nylon filter to remove particulates. Twenty microliters of filtered sample were fractionated on a high-performance liquid chromatography (HPLC) system with a C4 column (Waters XBridge Protein BEH, 2.1 mm x 250 mm, 300 Å pore size, 3.5 μm particle size). Proteins were separated at ambient temperature with a 0.2 mL/min flowrate using a linear gradient from 44% B to 60% B over 60 minutes (solvent A: 0.1% triflouroacetic acid:water; solvent B: 0.1% triflouroacetic acid:90% acetonitrile:water). Twenty 0.6 mL fractions were collected over the course of the gradient. Fractions were dried completely using a centrifugal vacuum concentrator. To reduce disulfide bridges, dried samples were reconstituted in liquid chromatography – mass spectrometry-grade water containing 20 mM tris(2-carboxyethyl)phosphine and incubated for 1 hour at ambient temperature with constant vortexing.

### Top-down analysis of proteins by mass spectrometry

Analyses were performed with a Bruker nanoElute (two-column method, Bruker TEN C18 column, 500 nL/min flow rate, 40⍰C) coupled with a Bruker timsTOF fleX mass spectrometer using the captive-spray source. Samples were first run in MS1-only mode (TIMS disabled, 1 Hz spectra rate, mass range 100-1700 m/z). Data was exported from the Bruker software in ASCII format. Using a custom MATLAB script, spectra were deconvoluted, and precursors were chosen for fragmentation. Samples were run again in auto-MS/MS mode with the chosen precursors added to the scheduled precursor list (TIMS disabled, 1 Hz MS spectra rate, 5 Hz MS/MS spectra rate, mass range 100-1700 m/z, active exclusion disabled, isolation width 2, collision energies ranging from 25-65 eV based on mass and charge state).

### Analysis of top-down proteomics data

Raw tandem mass spectrometry data were exported from the Bruker software in ASCII format. Spectra were deconvoluted and searched using custom MATLAB scripts developed in-house. An extensive proteoform database was created using neXtProt [22] SPARQL queries. The database included all human protein entries within the SwissProt database with a molecular weight ≤ 35 kDa (since DeepMALDI spectra were acquired in the range of 3,000 – 30,000 m/z). For each of these proteins, all allelic variants (with a frequency greater than 1%) and mature forms (e.g., signal- and pro-peptides removed) were also included. Truncations of up to three residues from either terminus were considered, as well as the following chemical modifications: water and ammonia loss, oxidation, hydroxylation, acetylation, succinylation, amidation, and specific glycosylations documented within the neXtProt database. Both MS and MS/MS data were utilized during matching, and scores for individual fragment ion matches were weighted based on the similarity between observed and predicted isotope distributions. Matches were confirmed by visual inspection of the spectra. Identified proteoforms were then mapped to the DeepMALDI spectra by comparison of their calculated masses to the m/z of the DeepMALDI peaks.

### Immunodepletion of Serum Amyloid A (SAA) and C-Reactive Protein (CRP) from Serum

Three microliters of VSP were diluted with 277 μL of PBS. 20 μL of biotin-conjugated monoclonal mouse anti-human SAA antibody were added to the diluted serum and incubated at room temperature for 2 hours. During the 2 hours of incubation, 100 μL of streptavidin modified magnetic particles (Dynabeads™ M-280 Streptavidin) were washed 3 times with a series of 900 μL, 100 μL and 100 μL of PBST. The diluted VSP and antibody mix was added to the washed Dynabeads and incubated for 30 minutes at room temperature. The antigen-antibody complex-coated Dynabeads were then separated from solution using the DynaMag™ −2 magnet (Thermo, Carlsbad, CA), and the resulting supernatant was transferred to a new tube. The supernatant from the antigen-antibody complex was mixed with 10 μL of biotin conjugated anti-SAA antibody and 100 μL of washed Dynabeads for a second round of SAA depletion following the same procedure described above. Ninety microliters out of the 300 μL of supernatant from the second round of depletion were transferred to a new tube for C18 clean-up and DeepMALDI analysis.

CRP was depleted from VSP using the procedure used for SAA depletion, with the following changes: 10 μL of biotin conjugated monoclonal mouse anti-human CRP antibody were added to the diluted serum and the second round of depletion was omitted.

Negative controls for the SAA and CRP depletions were performed in parallel following the exact same steps but without the addition of the anti-SAA and anti-CRP antibodies.

### C18 Cleanup of Immunodepleted Serum

C18 particles were activated with 50% acetonitrile:50% water and equilibrated with 0.5% trifluoroacetic acid in 5% acetonitrile. The immunodepleted serum and supernatant from corresponding controls (no antibody added) were brought up to a final concentration of 0.5% trifluoroacetic acid and 5% acetonitrile and mixed with the activated C18 particles in a spin filter tube. The unbound material was isolated by centrifugation. The flow-through collected by centrifugation was added back to the C18 particles for further binding. After the second centrifugation step, the C18 particles were washed with an aqueous solution of 0.5% trifluoroacetic acid in 5% acetonitrile. The C18-bound serum proteins were eluted with 2 × 20 μL of 70% acetonitrile in water. The eluents were combined, and the solvent removed with a centrifugal vacuum concentrator. The dried eluents were reconstituted in 20 μL of reagent grade water.

### Ethics statement

All samples were collected under ethics-approved protocols according to Institutional Review Board/Independent Ethics Committee requirements of the respective biobanks and the PROSE study (NCT00989690). Only anonymized patient samples were used, which were not obtained through investigator intervention or interaction with the individuals. The investigators cannot readily ascertain the identity of the individual(s) to whom the coded specimens pertain because there are agreements and IRB-approved policies and procedures in place that prohibit the release of the key to the code to the investigators under any circumstances until the individuals are deceased. Therefore, this research did not involve human subjects, and neither informed consent nor IRB review were required.

## Results

### Peak identification

The development of the VS algorithm [3] was based on the level of MALDI spectrometry available in 2005. Substantial progress has been made since then, both on the instrument and on the application side. Using DeepMALDI technology [2] it is now possible to measure proteoform abundance spanning a range of 4 orders of magnitude. We compared the peak content using this more sophisticated method with the original VS acquisition. The ability of the DeepMALDI method to elucidate the fine structure of peaks within the VS feature ranges can be seen in the overlay of VS and Deep MALDI spectra in the mass range relevant for VS in Figure 1.

**Figure 1.**
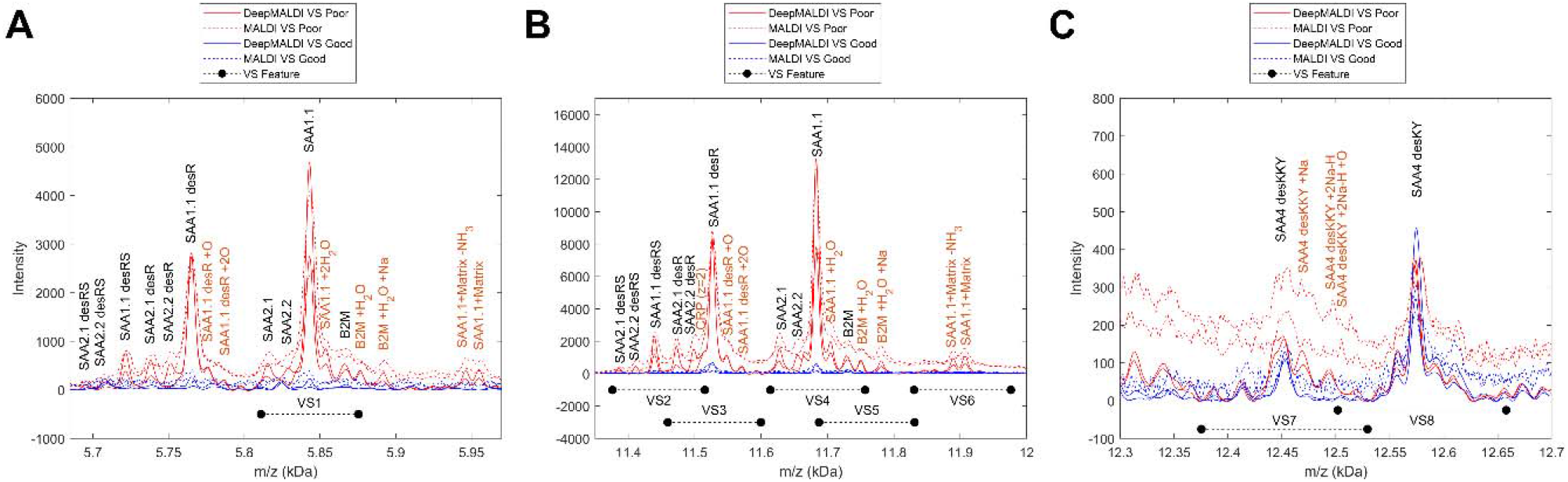
VS (dashed) and DeepMALDI (solid) spectra of two VS Poor (red) and two VS Good (blue) samples in the mass ranges relevant for VS. VS feature ranges are indicated below the spectra. Identified proteoform peaks are annotated in black for main proteoform peaks and in red for inferred proteoform + adducts peaks.

In the DeepMALDI spectra, we observe 81 distinct peaks within the 8 VS feature ranges. A listing of these peaks and their assignment to VS features is given in Supplementary Table 1. In order to gain a deeper understanding of the biological underpinnings of the VeriStrat test, we sought to identify the proteoforms that make up the VS features.

In a standard bottom-up proteomics approach, complex protein mixtures, such as serum, are digested with a protease, such as trypsin, prior to analysis by tandem mass spectrometry. Tandem mass spectra are used to identify peptides present in the digested mixture. These peptide identifications are then used to infer which protein groups were present in the original sample. This approach can lose proteoform-level information. Because DeepMALDI peaks represent intact proteoforms, it was necessary to identify the specific proteoforms present in serum samples. Thus, we performed top-down proteomics [23] in which whole proteoforms (no digestion) were introduced by liquid chromatography and electrospray ionization (ESI) onto the mass spectrometer for tandem mass spectrometric analysis.

Focusing on the predominant peaks in the VS spectra, we identified 44 proteoforms which constitute the major contribution to the values of the 8 VS features. These are listed in Table 2.

**Table 2.** A list of the 44 DeepMALDI peaks to which we were able to assign a proteoform descriptor using the notation as in [10]. The “des” nomenclature indicates that the proteoform is missing the indicated residues from one of its termini. For example, SAA4 desKKY is a proteoform of SAA4 in which the last three amino acid residues—KKY—are missing from the C-terminus. In the second column we show to which VS feature range a peak belongs. We have provided suitable references where available. The method of assignment is described in column 5. In the last column we indicate which peak identifications could be confirmed with immunodepletion experiments.

| m/z | VeriStrat Feature | Protein ID | References | Method of ID | Confirmed with Depletion |
| --- | --- | --- | --- | --- | --- |
| 5813 | VS1 | SAA2.1 (z=2) |  | Charge dependency |  |
| 5824 | VS1 | SAA2.2 (z=2) |  | Charge dependency |  |
| 5835 | VS2 | SAA2.2 +Na (z=2) |  | Charge dependency |  |
| 5842 | VS1 | SAA1.1 (z=2) |  | Charge dependency | Confirmed |
| 5861 | VS1 | SAA1.1 +2H<sub>2</sub>O (z=2) |  | Charge dependency | Confirmed |
| 5866 | VS1 | B2M (z=2) | [5] | Charge dependency |  |
| 5876 | VS1 | B2M +H<sub>2</sub>O (z=2) |  | Charge dependency |  |
| 11382 | VS2 | SAA2.1 desRS | [10, 24] | Top-down |  |
| 11408 | VS2 | SAA2.2 desRS | [10, 24] | Top-down |  |
| 11419 | VS2 | SAA1.1 desRS -NH<sub>3</sub> |  | Adducts inferred | Confirmed |
| 11436 | VS2 | SAA1.1 desRS | [10, 24] | Top-down | Confirmed |
| 11458 | VS2 | SAA1.1 desRS +Na |  | Adducts inferred | Confirmed |
| 11475 | VS2,VS3 | SAA2.1 desR | [10, 24] | Top-down |  |
| 11493 | VS2,VS3 | SAA2.2 desR | [10, 24] | Top-down |  |
| 11514 | VS2,VS3 | CRP (z=2) |  | Charge dependency | Confirmed |
| 11528 | VS3 | SAA1.1 desR | [10, 24] | Top-down | Confirmed |
| 11544 | VS3 | SAA1.1 desR + O |  | Adducts inferred | Confirmed |
| 11559 | VS3 | SAA1.1 desR +2O |  | Adducts inferred | Confirmed |
| 11627 | VS4 | SAA2.1 | [10, 24] | Top-down |  |
| 11648 | VS4 | SAA2.2 | [10, 24] | Top-down |  |
| 11669 | VS4 | SAA2.2 +Na |  | Adducts inferred |  |
| 11683 | VS4 | SAA1.1 | [10, 24] | Top-down | Confirmed |
| 11703 | VS4,VS5 | SAA1.1 +H<sub>2</sub>O |  | Adducts inferred |  |
| 11710 | VS4,VS5 | B2M -H<sub>2</sub>O |  | Adducts inferred |  |
| 11722 | VS4,VS5 | SAA1.1 +2H<sub>2</sub>O |  | Adducts inferred | Confirmed |
| 11730 | VS4,VS5 | B2M | [5] | Top-down |  |
| 11739 | VS4,VS5 | SAA1.1 +3H<sub>2</sub>O |  | Adducts inferred | Confirmed |
| 11748 | VS4,VS5 | B2M +H<sub>2</sub>O |  | Adducts inferred |  |
| 11762 | VS5 | B2M +H<sub>2</sub>O +O |  | Adducts inferred |  |
| 11778 | VS5 | B2M +H<sub>2</sub>O +2O |  | Adducts inferred |  |
| 11784 | VS5 | B2M +H<sub>2</sub>O +Na |  | Adducts inferred |  |
| 11893 | VS6 | SAA1+Matrix -NH<sub>3</sub> |  | Adducts inferred | Confirmed |
| 11906 | VS6 | SAA1+Matrix |  | Adducts inferred | Confirmed |
| 11936 | VS6 | B2M +Matrix -H<sub>2</sub>O |  | Adducts inferred |  |
| 11951 | VS6 | B2M +Matrix - H<sub>2</sub>O +O |  | Adducts inferred |  |
| 11963 | VS6 | SAA1.1 +Matrix +3x H<sub>2</sub>O |  | Adducts inferred |  |
| 11972 | VS6 | B2M +Matrix +H<sub>2</sub>O |  | Adducts inferred |  |
| 12443 | VS7 | SAA4 desKKY |  | Top-down |  |
| 12468 | VS7 | SAA4 desKKY +Na |  | Adducts inferred |  |
| 12490 | VS7 | SAA4 desKKY +2Na-H |  | Adducts inferred |  |
| 12505 | VS7,VS8 | SAA4 desKKY +2Na-H +O |  | Adducts inferred |  |
| 12527 | VS7, VS8 | SAA4 desKKY +3Na-2H +O |  | Adducts inferred |  |
| 12544 | VS8 | SAA4 desKKY +3Na-2H +O +NH<sub>3</sub> |  | Adducts inferred |  |
| 12572 | VS8 | SAA4 desKY |  | Top-down |  |

In terms of protein families, we identified serum amyloid A1 (SAA1, UniProt ID P0JI8), serum amyloid A2 (SAA2, UniProt ID: P0DJI9), serum amyloid A4 (SAA4, UniProt ID: P35542), beta-2-microglubulin (B2M, UniProt ID: P61769), and C-reactive protein (CRP, UniProt ID: P02741). Mass spectral peaks corresponding to the acute inflammatory proteins SAA1 and SAA2 and some of their isoforms had been previously identified in spectra of human serum/plasma [9, 10, 24]. B2M, as a marker of liver disease [5], and CRP, as a circulating protein associated with inflammatory processes and immune response, have also been identified in human serum or plasma spectra.

The association of inflammatory markers, in particular SAA and CRP, with VS classification, has also been reported previously [6, 25]. The association of SAA4 with VS features VS7 and VS8 is, to our knowledge, a novel discovery.

Confirming and extending upon a previous report [10], our use of top-down identification allowed for a direct assignment of different SAA alleles to different VS features. It is interesting to note that we observed the less common allele SAA2.2, allele frequency 0.140067, together with the more common allele SAA2.1, allele frequency 0.83539 [26], in a single patient’s sample (see Figures 1A/B).

DeepMALDI peaks in the lower mass range of the VS feature VS1 are doubly-charged peaks of the proteoforms in the higher mass VS features VS2 – VS6, as the pattern of peaks is identical, and the peak positions match nearly perfectly.

Additionally, in the DeepMALDI analysis we observed a well-defined peak at 11,514 Da, more visible in VS Good samples, which is absent in the doubly-charged VS1 range (see Figure 2). This peak was highly correlated (Pearson correlation of 0.92 in the PROSE set) with a peak at 23,045 Da, at the singly charged position. From the literature [27], we assigned this peak to CRP and confirmed by immunodepletion (Figure 3C). It is also not surprising that CRP cannot be observed in the VS1 range, because quadruply-charged ions are rare in MALDI when using sinapinic acid matrix [28].

**Figure 2.**
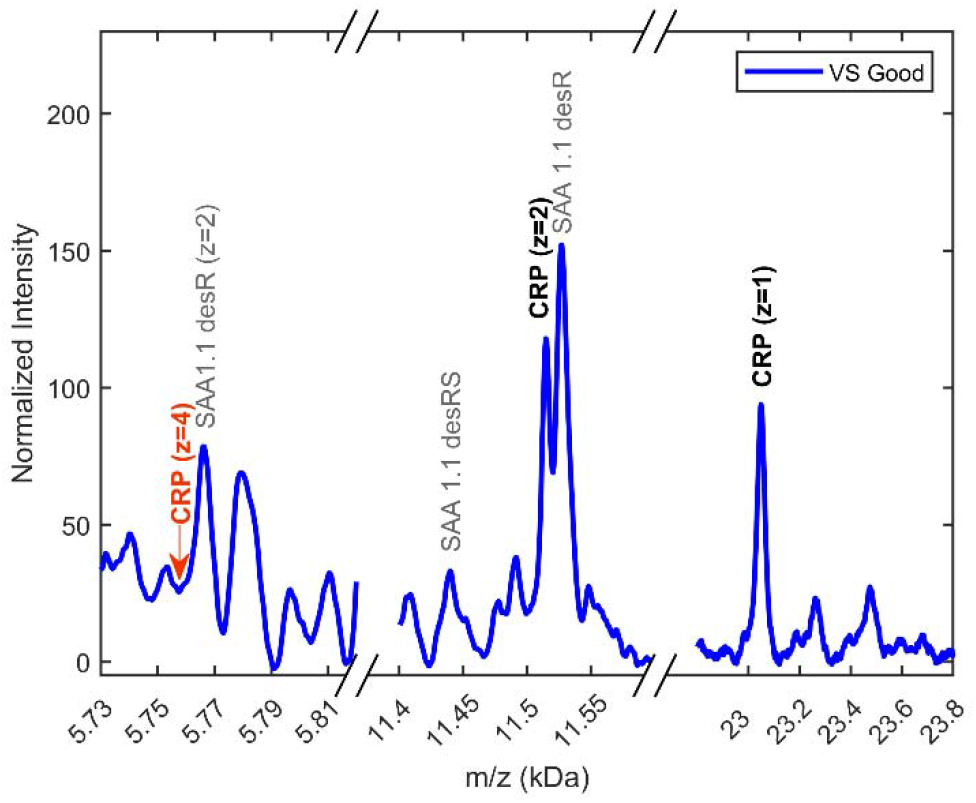
DeepMALDI spectrum of a VS Good sample over the mass ranges containing singly- and doubly-charged CRP and the range where one would expect a quadruply-charged CRP indicated by the red annotation “CRP (z=4)”.

**Figure 3.**
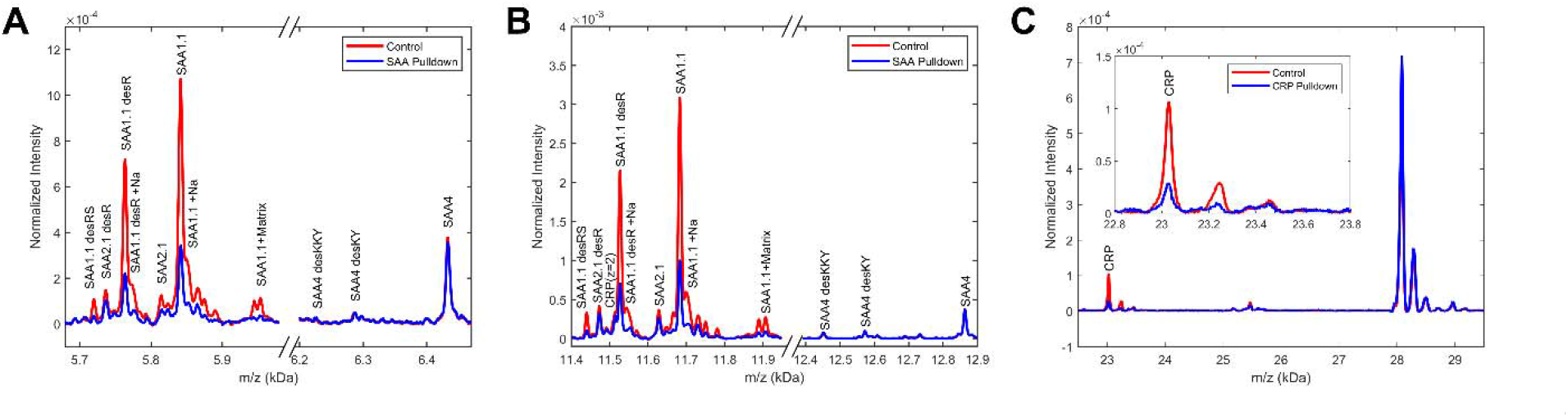
DeepMALDI mass spectra of a VS Poor sample (red) and the same sample after immunodepletion with an anti-SAA-antibody (A and B, for relevant singly- and doubly-charged mass ranges, respectively), and with a CRP antibody (C, in the singly-charged CRP range). The ranges have been extended to show that depletion did not largely affect other proteoforms.

The top-down procedure allowed for very good coverage of the protein fragments in the respective MS2 spectra allowing for the detailed identification of the observed proteoform. Examples of this matching procedure—and the high quality of the matches on both the MS1 and MS2 level—are shown for SAA4-desKY, B2M, and SAA1.1 (see supplementary Figures S1 – S3).

We performed selected depletion experiments to confirm top-down identifications with an orthogonal approach. In Figures 3A and 3B, we show the reduction in the SAA peaks due to depletion with an anti-SAA antibody. While almost all SAA peaks are reduced, it appears that the antibody used was more effective in binding to SAA1 than SAA2 (e.g. the reduction in the SAA1.1 peak is much more pronounced than that in the SAA2.1 peak). Additionally, SAA4 proteoforms were not significantly reduced, indicating specificity to the SAA1 and SAA2 isoforms. The reduction in peak height due to SAA immunodepletion also confirmed the assignment of peaks arising from adducts. The SAA1.1+matrix, SAA1.1+Na, and SAA1.1desR+Na peaks were strongly affected by depletion. As expected, the CRP(z=2) peak was not affected by this depletion.

### Correlation analysis of identified proteoforms

The association of inflammatory proteins, in particular SAA, with VS classification has been investigated in detail in other work [6, 25]. Here we advance this work by showing the association of the above identified DeepMALDI peaks with VS classifications and investigate the correlations of these peaks using as a data set the available samples from the PROSE study [11].

A heatmap showing the intensities of a subset (fewer adducts) of identified DeepMALDI peaks across individual VS Good and VS Poor samples is shown in Figure 4A. With the exception of the SAA4-desKY peaks, all other identified peaks are significantly associated with VS result (Mann-Whitney p <0.001, see supplementary Figure SF4). However, a definition of a single per-peak cut-off is difficult as there is a continuum of values for each feature, with overlap between VS groups in the low peak intensity range. Absence of simple cut-offs is not unexpected, as the VS test is multivariate in nature, with VS classification defined via a k-nearest neighbor (kNN) algorithm using a 26-sample reference set [3].

**Figure 4.**
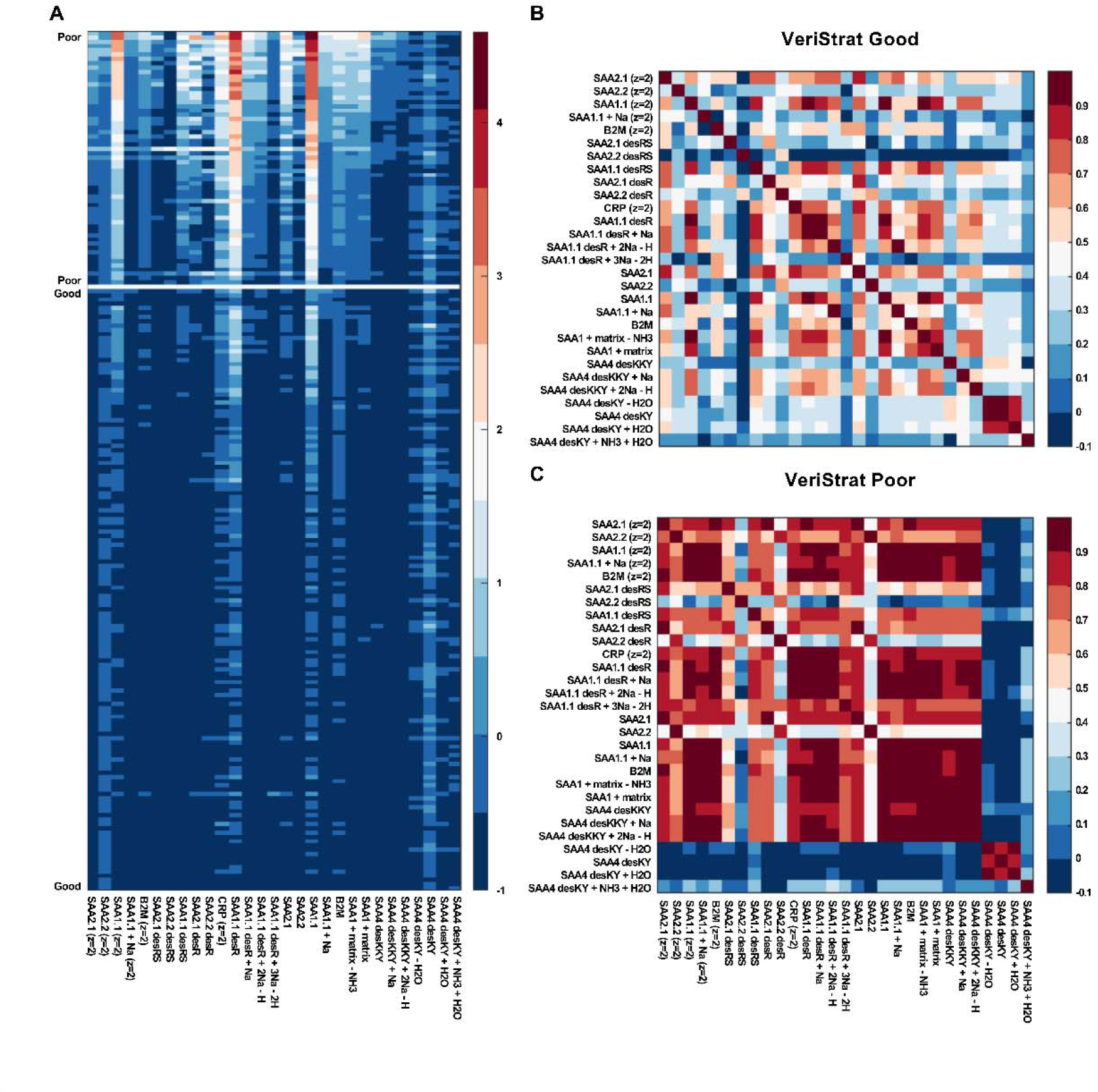
On the left, the peak intensities of the identified DeepMALDI peaks in the PROSE cohort (A). Intensity values were transformed using log(0.4 + intensity) to render smaller peaks visible in the presence of the large SAA1.1 peaks. As indicated on the left, on the top are the 59 VS Poor samples, and on the bottom are the 140 VS Good samples. Within each group, samples are in descending order of the value of SAA1.1 peak intensity. On the right, the Pearson correlations of these peaks are evaluated in the VS Good patients (B) and in the VS Poor patients (C).

From the Pearson correlations between identified proteoforms within in each VS group (see Figures 4B and 4C), additional evidence for our proteoform identification can be obtained. Off-diagonal correlations are in general larger for the VS Poor group than in the VS Good group. Peak intensities are generally higher in VS Poor samples, and we expect measurement reproducibility to be better for larger peaks. Since Pearson correlation is normalized by the standard deviation, it is not surprising that correlations are generally stronger within the VS Poor group. In the VS Good group correlations (Figure 4B), one can observe strong correlations between singly- and doubly-charged proteoforms and somewhat weaker associations of proteoforms with proteoforms+adducts. One can also observe a relatively strong correlation of all SAA1 peaks with one another and with CRP. This strongly correlated group extends to SAA2.1 and SAA4-desKKY in the VS Poor group correlations (Figure 4C). SAA2.2 is much less correlated with this group, likely due to its smaller allele frequency. SAA4-desKY forms a separate group, with the adduct SAA4-desKY+NH3+H20 only weakly correlated with it. It may be that the latter peak depends on sample pre-processing details, or that our inference of this SAA4 adduct peak based on mass matching is incorrect.

## Discussion

The blood-based VeriStrat proteomic test is based on eight MALDI-TOF spectral features spanning an m/z range of 5811 to 12658. As VS is based on Biodesix’s early generation MALDI-TOF technologies, each individual spectral feature is broad and has been thought to encompass multiple proteins and their proteoforms [6, 9, 10]. The latest iteration of DeepMALDI proteomics (circa 2021) was used to discover the fine structure spectrum underlying each of the VS features, finding that the eight VS features are composed of 81 underlying protein peaks. A combination of top-down proteomics and inferences based on mass were used to identify the proteoforms responsible for 44 of these peaks, and immunodepletion was used to validate many of these findings. We found that CRP and B2M, as well as different proteoforms of SAA1, SAA2, and SAA4 make up these 44 spectral peaks and are thereby the major constituents of the eight VS features.

The VeriStrat proteomic test has recently been shown in the INSIGHT clinical trial [1] and by Chae *et al* [16] to identify those patients with non-small cell lung cancer most likely to respond to immune checkpoint inhibitors. There is indication that the test may be useful for determining those patients that would best benefit from the addition of platinum doublet chemotherapy to ICI therapy. One of the goals of definitively identifying the VS proteins in this present study was to better understand how they may play a role in the immunobiology of cancer, and hence as relevant biomarkers of response to ICI treatments.

A number of studies have examined SAA, CRP, and B2M individually, as univariate biomarkers, and as potential prognostic and predictive biomarkers of various cancer outcomes [29-35]. Additional works have examined these proteins as potential predictive biomarkers of outcomes following ICI therapy in different cancers including lung, breast, colon, melanoma, and others [30, 32, 33, 35, 36]. Yet, none of these studies have resulted in these proteins being used as individual clinical biomarkers for cancer, most likely due to the inherent problems of applying univariate analyses to covariance and correlation in complex biologic systems, and using small non-representative patient cohorts for biomarker discovery [37-40]. For example, a review and meta-analysis of CRP found that, among 33 studies, 19 different cutoff levels were identified for CRP to be prognostic of ICI outcomes [41]. This is consistent with our finding that at the decision margin of the VS test there is considerable overlap between the abundance of these markers, which renders the definition of a simple threshold difficult.

On the other hand, by measuring multiple features related to an outcome, multivariate tests address the inherent problems of univariate tests [42, 43]. The VeriStrat test takes advantage of the increased fidelity of multivariate analysis by incorporating SAA isoforms, CRP, and B2M into one test that has been repeatedly validated in both prognostic and predictive settings for NSCLC outcomes [29-35].

Most recently, the VeriStrat test has been shown to stratify outcome (VS groups) for patients receiving single agent immunotherapy and triplet therapy [1], with point-estimates showing a differential outcome. A question raised by this finding is: *How are the proteins measured by the VeriStrat test related to immunotherapy outcomes from a prognostic and a biologic perspective?*

SAA proteins increase expression up to 1000-fold during the acute phase response, which is part of an attempt to restore homeostasis after an immune response or tissue insult [44]. SAA is prognostic of outcomes in NSCLC and other cancers [29, 30, 32], as well as efficacy and outcomes of ICI therapy [45, 46]. One important point to note is that it is not yet determined how the different SAA proteins and their proteoforms individually have roles in prognosis or prediction. SAA has been shown to activate myeloid derived suppressor cells that inhibit the immune system [47, 48]. Therefore, increased SAA may have a direct effect on the efficacy of ICIs by acting to suppress the immune system even when PD1/PD-L1 is inhibited.

CRP is the prototypic acute phase reactant protein [49] with increased expression during inflammation and tissue damage [50]. CRP is a recognized biomarker of inflammation [51, 52] that is routinely used as a clinical biomarker for overall immune system activity. Numerous studies have demonstrated the potential of CRP as prognostic for outcomes of ICI therapy of cancers [41]. Yet, as described above, a definitive cutoff level for CRP has not been established to make it clinically useful in the ICI context. Interestingly, CRP may play an active role in dampening the adaptive immune system [53], lending the possibility that it may suppress the immune system even in the context of PD1/PD-L1 inhibition.

B2M is critical for presentation of antigens, including potential tumor antigens by tumor cells, in the context of the major histocompatibility complex class 1 complex (MHC I) [54, 55]. B2M is required for loading antigen into MHC I. As cytotoxic CD8+ T cells recognize MHC I-bound antigen on the surface of cells, B2M is critical for CD8+ binding to and lysis/killing of tumor cells [56]. Elevated levels of B2M in blood have been shown in the Atherosclerosis Risk in Communities (ARIC) study to be associated with increased risk of cancer development, including NSCLC [57, 58], and elevated in different cancers [59, 60]. Differing levels of B2M have been shown to be prognostic of outcomes in various cancers [61-64]. Additionally, B2M has been shown to correlate with ICI therapeutic response in NSCLC and melanoma [65, 66], likely through disruption of tumor antigen presentation [67].

As noted above, the VeriStrat proteomic test has recently been shown to identify patients with NSCLC that are most likely to respond to ICI therapy. There are hints that it may assist in answering the question who would receive most benefit from the addition of platinum doublet therapy to ICI[1, 16]. In this context, we propose a model (Figure 5) where the proteins measured by the VeriStrat proteomic test are active players in the anti-tumor immune response. Namely that increased levels of SAA and CRP are immunosuppressive, and differing levels of B2M expression likely change tumor antigen presentation status. These immuno-oncology effects are a possible explanation for why the increased levels of SAA and CRP, and changes in B2M levels, contribute to suppression of ICI activity as well as to why the VeriStrat proteomic test is predictive of ICI treatment outcomes.

**Figure 5.**
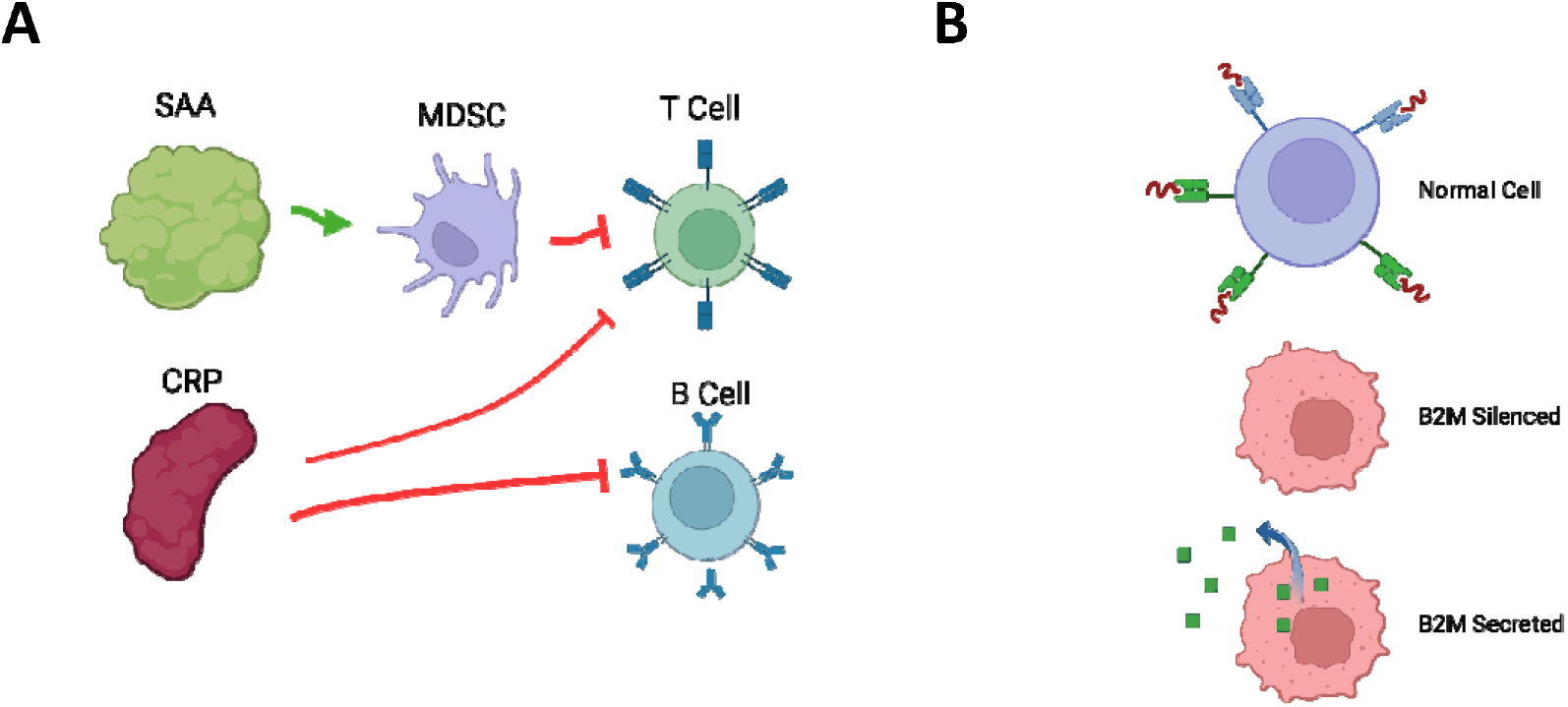
Proposed immunomodulatory actions of SAA, CRP, and B2M in the context of the anti-tumor immune response. (A) Increased levels of CRP and SAA proteins have been shown to suppress the adaptive immune system. SAA activates MDSC which suppress T cell activity. CRP has been shown to suppress both T cell and B cell activity. (B) Normal cells express normal levels B2M which interacts with MHC class I to allow antigen loading and display on the cell surface to enable recognition by cytotoxic T cells. Cancer cells can silence expression of B2M through a number of mechanisms resulting in a loss of MHC class I antigen presentation. Alternatively, cancer cells can export B2M to the extracellular space preventing its interaction with MHC class I, again resulting in loss of antigen presentation.

## Conclusions

This study has identified B2M and CRP, as well as multiple proteoforms of SAA1, SAA2, and SAA4 as the predominant proteins measured by the VeriStrat proteomic test. These proteins have been shown to have prognostic value for various cancer outcomes. Also, they all play active roles in cancer immunobiology leading to a possible mechanistic explanation as to the VeriStrat proteomic test’s ability to predict those patients that may or may not benefit from the addition of chemotherapy to ICI treatment.

## Data Availability

Data produced in the present study are available upon reasonable request to the authors.

## Abbreviations

B2M: beta-2 microglobulin
CRP: C-reactive protein
ESI: electrospray ionization
HPLC: high-performance liquid chromatography
ICI: immune checkpoint inhibitor
IGF-2: insulin-like growth factor 2
IL-6: interleukin-6
kNN: k-nearest neighbors
LC-MS/MS: liquid chromatography – tandem mass spectrometry
MALDI: matrix-assisted laser desorption/ionization
MHC I: major histocompatibility complex class I
NSCLC: non-small cell lung cancer
PBS: phosphate-buffered saline
PBST: PBS Tween
SAA: serum amyloid A
TOF: time-of-flight
VS: VeriStrat
VSG: VS Good serum pool
VSP: VS Poor serum pool

## CRediT author statement

**Matthew A Koc:** Methodology, Software, Formal analysis, Writing – Original Draft, Writing – Review & Editing, Visualization **Timothy Aaron Wiles:** Methodology, Investigation, Writing – Original Draft, Writing – Review & Editing, Visualization **Dan Weinhold:** Investigation, Writing – Review & Editing **Steven Rightmyer:** Investigation, Writing – Review & Editing **Joanna Roder:** Conceptualization, Writing – Original Draft, Writing – Review & Editing **Robert W Georgantas III:** Conceptualization, Writing – Original Draft, Writing – Review & Editing, Project administration **Senait Asmellash:** Conceptualization, Methodology, Writing – Original Draft, Writing – Review & Editing, Supervision **Heinrich Roder:** Conceptualization, Methodology, Writing – Original Draft, Writing – Review & Editing, Visualization, Supervision, Project administration

## Acknowledgements

The authors thank Amanda Weaver (Development, Biodesix INC, Boulder, CO) for collecting VS spectra on the Bruker microflex instrument.

## Data availability

Data produced in the present study are available upon reasonable request to the authors.

## Funding

This research did not receive any specific grant from funding agencies in the public, commercial, or not-for-profit sectors.

## Declaration of Competing Interest

The authors declare the following financial interests/personal relationships which may be considered as potential competing interests: All authors are current employees of and have or had stock options in Biodesix, Inc. H.R., J.R., and S.A. are inventors on patents describing DeepMALDI, assigned to Biodesix, Inc.

## Notes

### Author Declarations

All samples were collected under ethics-approved protocols according to Institutional Review Board/Independent Ethics Committee requirements of the respective commercial biobanks - Discovery Life Sciences (Huntsville, AL), ProMedDx (Norton, MA), ProteoGenex (Inglewood, CA) and BioIVT (Westbury, NY) - and the PROSE study (NCT00989690). Only anonymized patient samples were used, which were not obtained through investigator intervention or interaction with the individuals. The investigators cannot readily ascertain the identity of the individual(s) to whom the coded specimens pertain because there are agreements and IRB-approved policies and procedures in place that prohibit the release of the key to the code to the investigators under any circumstances until the individuals are deceased. Therefore, this research did not involve human subjects, and neither informed consent nor IRB review were required.

