## Supplementary Figures and Tables for "Molecular & Translational Biology of the Blood-Based VeriStrat® Proteomic Test Used in Cancer Immunotherapy Treatment Guidance"

### Supplementary Materials

**Table S1.** A list of m/z values for all DeepMALDI proteomic method peaks within the VS feature ranges. Some of the VS features overlap; thus, some peaks belong to more than one feature.

| **VS1** | **VS2** | **VS3** | **VS4** | **VS5** | **VS6** | **VS7** | **VS8** |
| --- | --- | --- | --- | --- | --- | --- | --- |
| 5810 | 11372 | 11465 | 11617 | 11696 | 11848 | 12383 | 12505 |
| 5813 | 11382 | 11475 | 11627 | 11703 | 11861 | 12397 | 12516 |
| 5824 | 11391 | 11493 | 11648 | 11710 | 11869 | 12410 | 12527 |
| 5829 | 11408 | 11502 | 11655 | 11722 | 11876 | 12430 | 12544 |
| 5835 | 11419 | 11514 | 11669 | 11730 | 11893 | 12443 | 12556 |
| 5842 | 11427 | 11528 | 11683 | 11739 | 11906 | 12449 | 12572 |
| 5861 | 11436 | 11544 | 11696 | 11748 | 11920 | 12468 | 12591 |
| 5866 | 11444 | 11552 | 11703 | 11762 | 11936 | 12482 | 12608 |
| 5876 | 11458 | 11559 | 11710 | 11778 | 11951 | 12490 | 12621 |
|  | 11465 | 11571 | 11722 | 11784 | 11963 | 12505 | 12631 |
|  | 11475 | 11580 | 11730 | 11790 | 11972 | 12516 | 12654 |
|  | 11493 | 11593 | 11739 | 11805 |  | 12527 |  |
|  | 11502 |  | 11748 | 11816 |  |  |  |
|  | 11514 |  |  | 11829 |  |  |  |

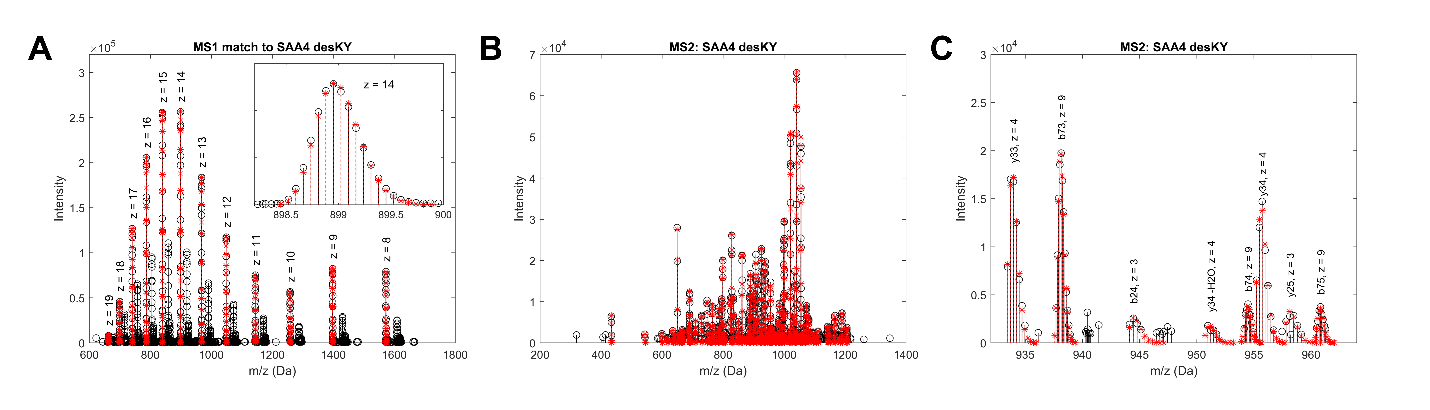

**Figure S1.** Pooled human serum was fractionated on a C4 column, and fractions were reduced to break disulfide bridges. Reduced fractions were analyzed by liquid chromatography – tandem mass spectrometry. Data were searched against an extensive proteoform database using a MATLAB script developed in-house. **(A)** MS1 spectrum showing multiple charge states of a precursor that was matched to the proteoform SAA4 desKY. Observed peaks are shown in black and predicted peaks are shown in red. For each isotope cluster, the theoretical distribution was scaled to fit the observed distribution using a least-squares approach. The inset shows the close match between the predicted and observed isotope distribution for one of the charge states. **(B)** Observed MS2 spectrum for the isolated precursor annotated with predicted isotope distributions of theoretical fragment ions for the SAA4 desKY sequence. **(C)** Visual inspection of a smaller m/z range shows nice fits between predicted and observed b- and y-ions.

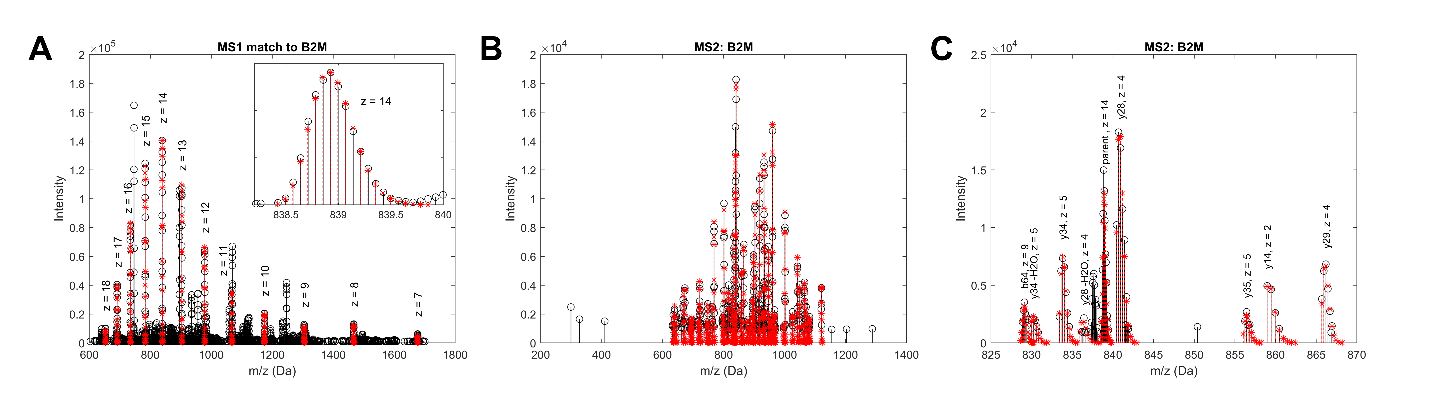

**Figure S2.** Pooled human serum was fractionated on a C4 column, and fractions were reduced to break disulfide bridges. Reduced fractions were analyzed by liquid chromatography – tandem mass spectrometry. Data were searched against an extensive proteoform database using a MATLAB script developed in-house. **(A)** MS1 spectrum showing multiple charge states of a precursor that was matched to the proteoform B2M. Observed peaks are shown in black and predicted peaks are shown in red. For each isotope cluster, the theoretical distribution was scaled to fit the observed distribution using a least-squares approach. The inset shows the close match between the predicted and observed isotope distribution for one of the charge states. **(B)** Observed MS2 spectrum for the isolated precursor annotated with predicted isotope distributions of theoretical fragment ions for the B2M sequence. **(C)** Visual inspection of a smaller m/z range shows nice fits between predicted and observed b- and y-ions.

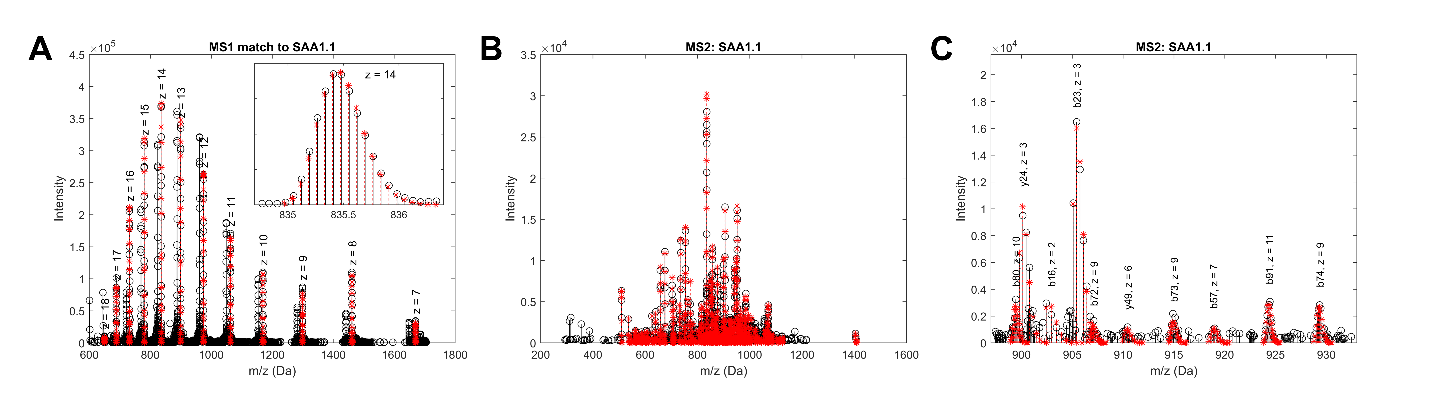

**Figure S3.** Pooled human serum was fractionated on a C4 column, and fractions were reduced to break disulfide bridges. Reduced fractions were analyzed by liquid chromatography – tandem mass spectrometry. Data were searched against an extensive proteoform database using a MATLAB script developed in-house. **(A)** MS1 spectrum showing multiple charge states of a precursor that was matched to the proteoform SAA1.1. Observed peaks are shown in black and predicted peaks are shown in red. For each isotope cluster, the theoretical distribution was scaled to fit the observed distribution using a least-squares approach. The inset shows the close match between the predicted and observed isotope distribution for one of the charge states. **(B)** Observed MS2 spectrum for the isolated precursor annotated with predicted isotope distributions of theoretical fragment ions for the SAA1.1 sequence. **(C)** Visual inspection of a smaller m/z range shows nice fits between predicted and observed b- and y-ions.

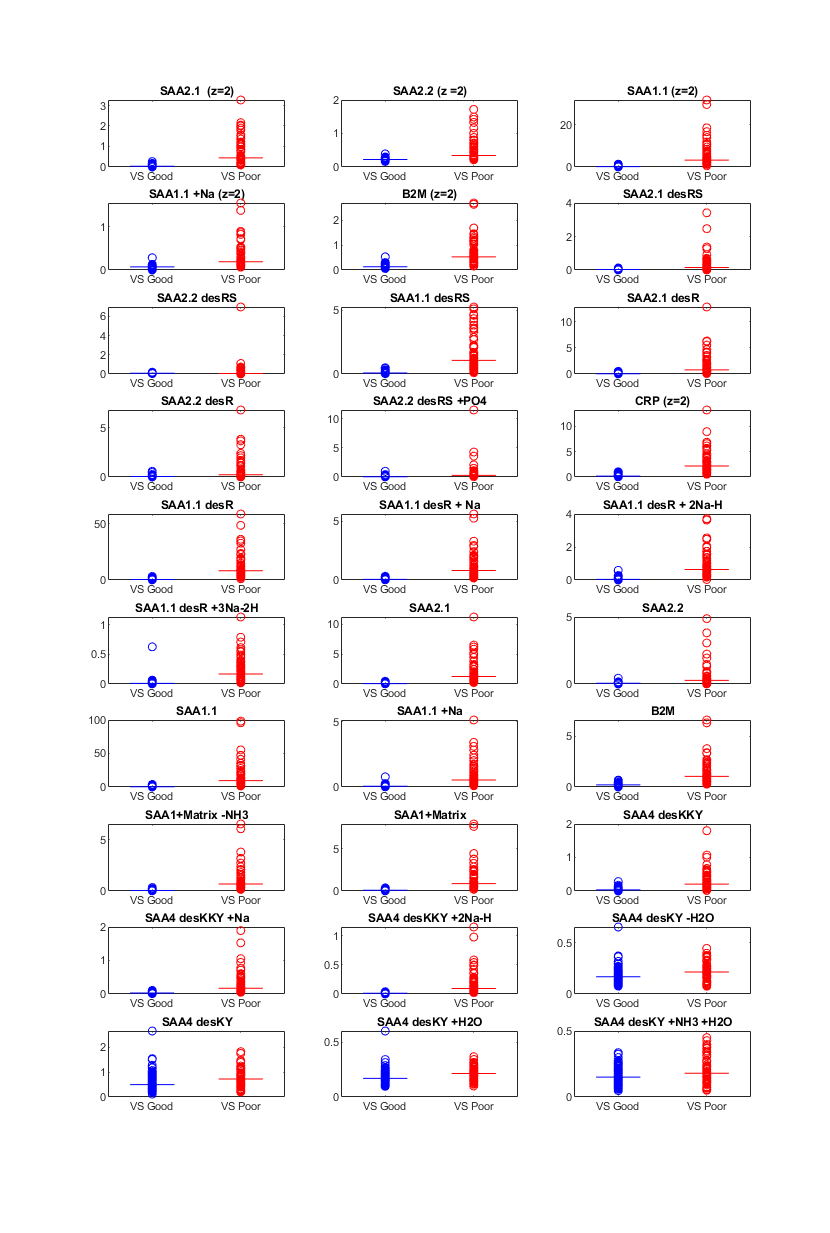

**Figure S4.** Distribution of peak intensity of identified proteoforms by VS group (VS Good in blue and VS Poor in red) for the PROSE set. Medians per VS group are shown as horizontal lines.
